# A Vision-Language Framework for Predicting Brain Tumor Recurrence from Multimodal, Longitudinal Patient Data

**DOI:** 10.64898/2026.08.11.26360196

**Authors:** Divyanshu Tak, Diya Sreedhar, Hugo JWL Aerts, Benjamin H Kann

## Abstract

Accurate prediction of tumor recurrence in brain tumor patients following surgery is essential for optimizing adjuvant therapy, response assessment, and surveillance regimen. While MRI remains the gold standard for surveillance, integrating patient-specific clinical context may inform recurrence prediction. Traditional multimodal deep learning approaches often incorporate clinical data via simple fusion, failing to fully capture the semantic interdependencies between visual features and clinical context. Trained on over 5,000 scans from approximately 400 pediatric low-grade glioma subjects and validated across three institutional cohorts, including one clinical trial cohort, our experiments demonstrate incremental performance gains when progressing from vision-only to clinical-vision to a vision-language approach. Our results indicate that converting structured clinical covariates into natural language text allows for more effective synthesis of multimodal data, while providing a platform for incremental addition of clinical context without extending model complexity. We demonstrate that our proposed VLM architecture offers a promising direction for neuro-oncological prognosis by effectively encoding imaging cues and clinical context, with potential applicability to other longitudinal prognosis tasks.

## 1 Introduction

Pediatric brain tumors are the leading cause of cancer-related mortality in children, accounting for approximately 20% of childhood cancers. For patients with low-grade gliomas (LGGs), nearly 35% of all pediatric brain tumors, surgical resection followed by longitudinal MRI surveillance remains the standard of care[1-3]. However, clinical outcomes are highly heterogeneous: while many patients achieve long-term control, up to 50% experience recurrence requiring additional interventions[4,5]. By contrast, pediatric high-grade gliomas (HGGs) exhibit aggressive growth and worse outcomes, necessitating an even greater need for prognostic modeling. Accurate, early recurrence prediction could enable personalized surveillance protocols and timely therapeutic interventions that prevent clinical symptoms and complications of recurrence.

This clinical motivation has spurred increasing interest in AI-driven prognostic modeling[4, 6-9], yet existing approaches remain limited in how they integrate multimodal patient information leading to suboptimal performance and generalizability that are barrier to clinical translation[8-11]. Traditional multimodal deep learning frameworks typically encode clinical variables as tabular features concatenated with imaging representations, treating clinical data as static numerical inputs rather than contextual information[2,7,8,12]. This approach does not capture the semantic relationships between radiographic features and patient-specific clinical presentation, including demographics, surgical resection status, treatment intervals. In addition, data are generally analyzed as static timepoints, which results in information loss given the longitudinal nature of patient care and surveillance. Furthermore, the broader deployment of current AI prognosis tools for brain tumor recurrence is hindered by a scarcity of external validation on heterogeneous cohorts. This failure to demonstrate generalizability across different demographics remains a primary bottleneck for clinical translation [4,5,9].

### Related Work

Longitudinal Clinical AI models predominantly focus on capturing temporal dynamics from serial imaging using RNNs, transformers, or attention modules, rarely including patient-specific clinical context[2,7,8,13-16]. While multimodal fusion strategies attempt to bridge this gap by incorporating tabular data (e.g., demographics), they typically treat clinical variables as static fixed vectors rather than contextual conditioning information that semantically interacts with visual features[2,7,8,12]. Recently, Vision-Language Models (VLMs) have shown promise in aligning imaging with free-text reports and clinical context via cross-modal attention[13, 17-19]. However, existing medical VLMs are largely restricted in terms of limited longitudinal span (<=2 timepoints) [17-19] and VLM applications in recurrence prediction, with extended longitudinal timespans remains unexplored [4,8,20-22].

### Contribution

In this work, we introduce the first Vision-Language Model (VLM) framework for longitudinal prediction of tumor recurrence in pediatric brain tumor patients. Our approach converts clinical variables into structured natural language prompts processed through cross-attention mechanisms alongside longitudinal MRI sequences (Figure 1). We compare three strategies: vision-only approach, standard clinical-vision fusion with structured clinical variables, and our VLM approach(Figure 2). Trained on over 5,000 MRI scans from approximately 427 patients and validated across four independent institutional cohorts, including a completed clinical trial (PNOC008), our results demonstrate incremental gains in predictive performance with vision-language integration (Figure 3)[4,8,20]. This framework enables multimodal synthesis and allows expansion of clinical context without increasing model complexity. Our framework offers a robust approach for predictive longitudinal prognosis tasks in neuro-oncological and other biomedical longitudinal applications.

**Fig. 1.**
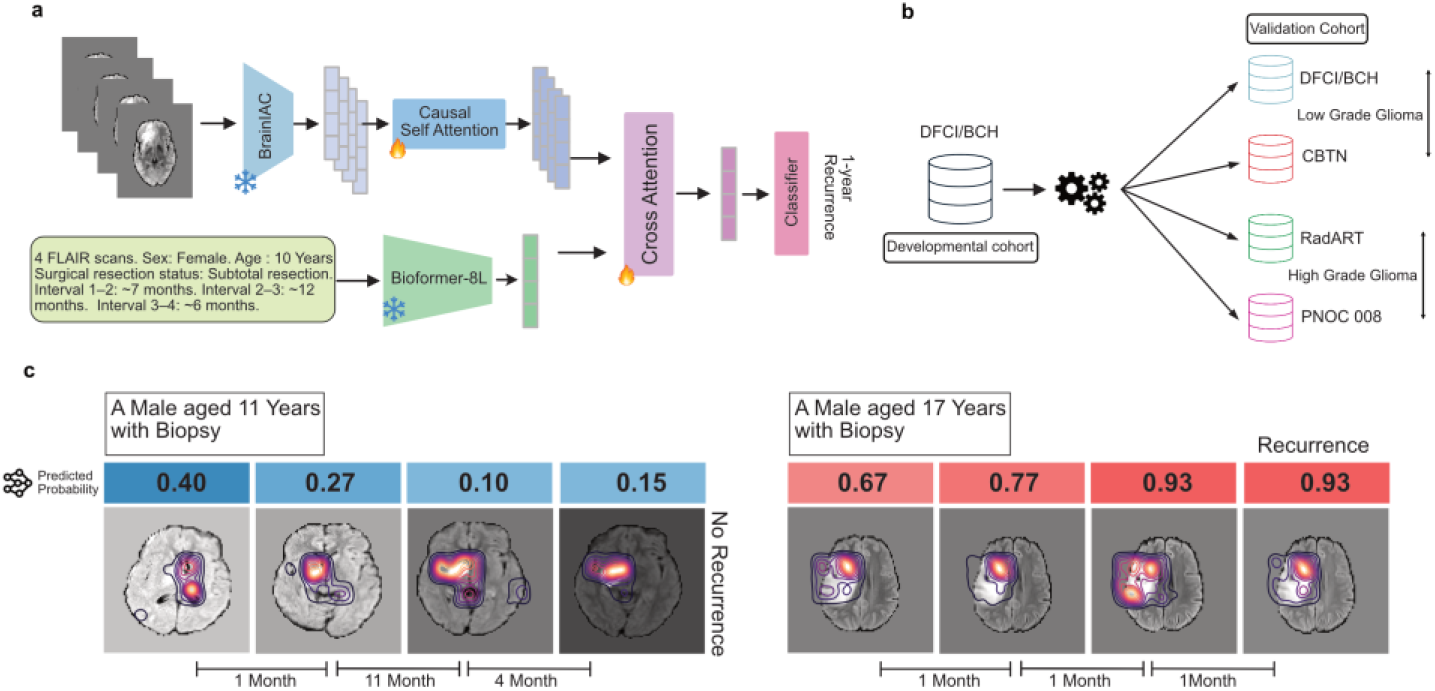
Overview of the proposed longitudinal vision–language modeling framework and external evaluation. (a) A sequence of FLAIR MRI scans is encoded with the frozen BrainIAC backbone and causal self-attention to allow temporal correlation, while clinical variables and inter-scan time intervals are expressed as structured natural-language prompts and embedded using a frozen Bioformer-8L encoder; cross-attention is used to fuse imaging and text representations, and a final classifier outputs 1-year recurrence risk. (b) The model is developed on a developmental cohort from DFCI/BCH and evaluated across four external sets covering both low-grade (DFCI/BCH, CBTN) and high-grade (RadART, PNOC008) gliomas to assess institutional and pathology based generalization. (c) Longitudinal examples illustrate predicted recurrence probabilities of the VLM and corresponding saliency maps across multiple surveillance time points, demonstrating stable low-risk trajectories in non-recurrence cases and progressively increasing risk preceding recurrence in high-risk cases.

**Fig. 2.**
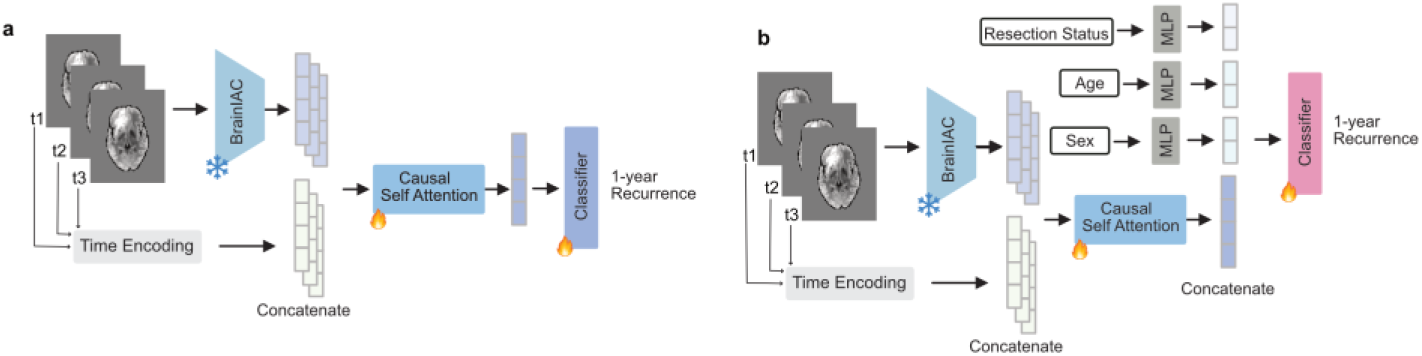
Architectures for Vision-only and Clinical-vision models. (a) **Vision-only model:** Each longitudinal MRI scan is encoded using the frozen BrainIAC backbone, while inter-scan time are converted into embeddings and concatenated with imaging tokens. A causal self-attention module processes the temporally ordered sequence, enforcing uni-directional information flow, and a final classifier predicts 1-year recurrence risk. (b) **Clinical-vision model:** In addition to temporal embeddings and imaging features, clinical covariates (resection status, age, sex) are encoded by MLP and fused with the longitudinal imaging representation before classification.

**Fig. 3.**
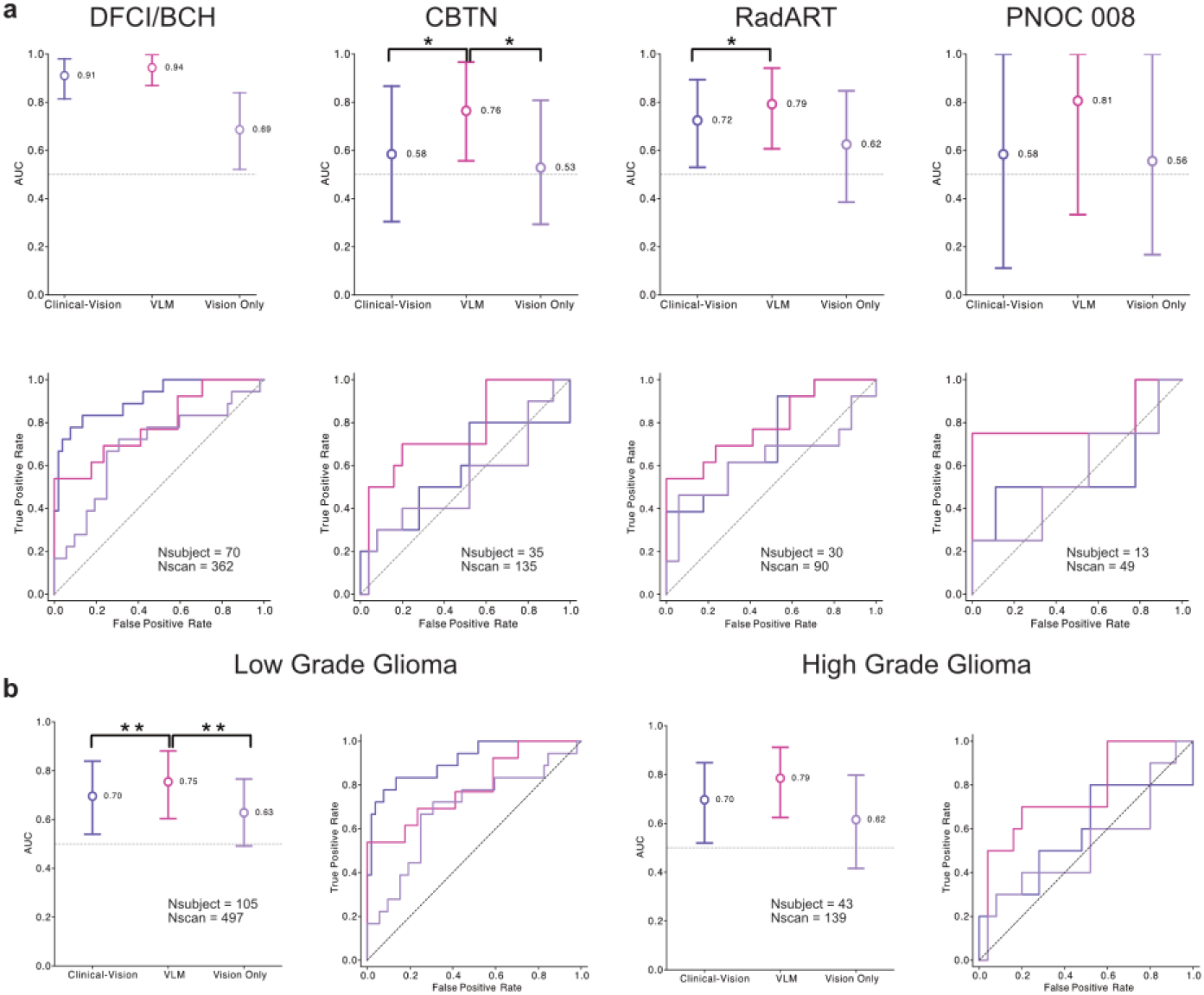
Model performance across external cohorts and pooled disease-grade analyses. (a) Comparison of the vision-only, clinical-vision, and proposed VLM model across four independent external cohorts (DFCI/BCH, CBTN, RadART, PNOC008). The VLM consistently outper-formed other models, with highest performance delta in the CBTN and PNOC008 cohorts. (b) Pooled analyses stratified by disease grade demonstrate similar patterns: in low-grade gliomas (N=105 subjects, 497 scans), the VLM substantially outperforms both baselines, while in high-grade gliomas (N=43 subjects, 139 scans), VLM maintains improved performance.

**Fig. 4.**
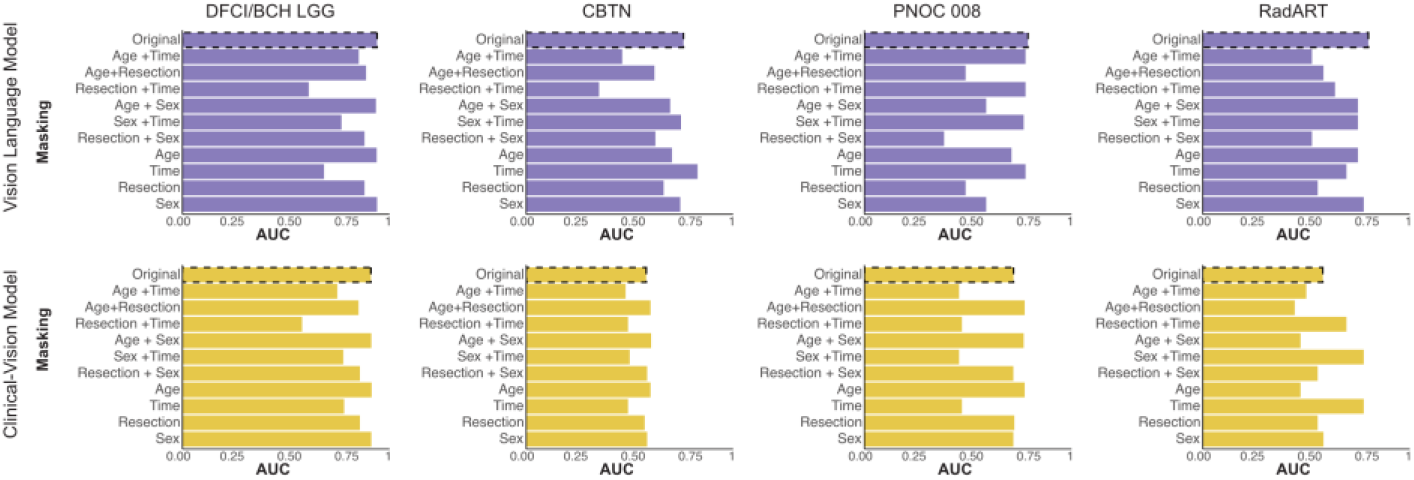
Ablation analysis of clinical variable contributions in recurrence prediction. Structured masking inference sweeps were performed to assess the influence of individual and pair-wise clinical attributes (age, sex, resection status, and inter-scan time intervals) on predictive performance for both the Vision Language Model (top row) and the clinical-vision baseline (bottom row) across all four external cohorts (DFCI/BCH, CBTN, PNOC008, RadART),

## 2 Methods

### 2.1 Data Cohort and Longitudinal Imaging data

We retrospectively collected longitudinal MRI surveillance scans from pediatric patients with pathologically confirmed low-grade glioma (LGG) treated across DanaFarber Cancer Institute-Boston Childrens Hospital (DFCI/BCH), Children’s Hospital of Philadelphia (CBTN), and University of California San-Francisco Medical Center (RadART). The high-grade glioma (HGG) data was collected from one multi-institutional clinical trial cohort (PNOC008). PNOC008 [23] is a pilot study assessing the clinical benefit of using molecular profiling to determine individualized treatment plans for pediatric and young adult patients with newly diagnosed HGG (NCT03739372). The final dataset comprised more than 5,000 MRI scans from 390 subjects. All datasets included T2-FLAIR MRI of patients who underwent primary surgery and had at least 1 year of clinical and radiographic follow-up. All postoperative MRI scans were included up until 1 year (365 days) prior to last clinical followup and/or diagnosis of recurrence, whichever came first (Table 1). An event was defined by radiology report impression of recurrence and/or progression, new clinical symptom attributable to tumor, change in patient management, and/or death per retrospective record review.

**Table 1.** Summary of demographic, clinical, and longitudinal imaging characteristics across the PNOC, CBTN, BCH-LGG, and RADART cohorts..

| Variable | BCH-LGG | CBTN | PNOC | RADART |  |
| --- | --- | --- | --- | --- | --- |
| N (subjects) | 312 | 35 | 30 | 13 |  |
| **Age at diagnosis, years** |  |  |  |  | **Age at diagnosis, years** |
| Mean ± SD | 9.1±5.4 | 8.2±5.7 | 10.5±4.7 | 13.2±4.8 | |
| Median (range) | 8 (0-20) | 7 (0-19) | 11 (2-18) | 13 (7-23) |  |
| **Sex n(%)** |  |  |  |  | **Sex, n (%)** |
| Male | 159 (51%) | 42 (58%) | 14 (47%) | 10 (77%) |  |
| Female | 153 (49%) | 31 (42%) | 16 (53%) | 3 (23%) |  |
| **Recurrence status, n (%)** |  |  |  |  | **Recurrence status, n (%)** |
| Recurrence | 139 (45%) | 34 (47%) | 13 (43%) | 4 (31%) |  |
| Non-recurrence | 173 (55%) | 39 (53%) | 17 (57%) | 9 (69%) |  |
| **Surgical resection status, n (%)** |  |  |  |  | **Surgical resection status, n (%)** |
| Biopsy | 66 (21%) | 34 (47%) | 16 (53%) | 1 (8%) |  |
| Gross total resection | 95 (30%) | 4 (5%) | 9 (30%) | 6 (46%) |  |
| Subtotal resection | 122 (39%) | 35 (48%) | 5 (17%) | 0 (0%) |  |
| Near total resection | 35 (48%) | 0 (0%) | 0 (0%) | 1 (8%) |  |
| Partial resection | 0 (0%) | 0 (0%) | 0 (0%) | 5 (38%) |  |
| **Number of scans per patient** |  |  |  |  | **Number of scans per patient** |
| Mean ± SD | 5.2±1.3 | 3.4±1.3 | 3.0±1.5 | 3.8±1.2 | |
| Range | 2-6 | 2-5 | 1-5 | 2-5 |  |
| **Average scan interval (months)** |  |  |  |  | **Average scan interval (months)** |
| Mean ± SD | 14.1±11.6 | 10.8±8.9 | 4.0±2.5 | 19.4±13.1 | |
| **Total follow-up interval (months)** |  |  |  |  | **Total follow-up interval (months)** |
| Mean ± SD | 62.1±58.4 | 27.7±28.3 | 10.1±10.5 | 55.5±45.9 | |

### 2.3 Dataset Preprocessing and Prompt Construction

T2 FLAIR MRI sequences were used as the primary imaging modality as they are routinely acquired in clinical surveillance and are the primarily recommended sequence for gauging treatment response and tumor recurrence in pediatric gliomas [1,3,25]. The preprocessing pipeline consisted of DICOM-to-NIfTI conversion, N4 bias field correction, skull stripping. For each subject, the longitudinal trajectory was represented as an ordered sequence of post-operative scans {s_1_,s_2_,…,s_7_}, with a maximum of seven scans retained to ensure uniform modeling across patients[4,7,8].

Clinical variables were extracted from retrospective clinical database and trial registries,including demographic features such as age and sex, surgical resection status, which is the clinical feature most associated with recurrence [REF], and the dates of surgery and MRI surveillance scans. Recurrence labels were assigned based on documented radiographic or clinical progression occurring within one year after the final scan in the modeled trajectory.

To integrate clinical context and temporal information, each trajectory was paired with a structured text prompt. The prompt summarized (i) the number of scans in the trajectory, (ii) clinical characteristics and (iii) the inter-scan intervals in months. A representative template of the prompt format is shown:

> *“4 FLAIR scans. Sex: Female. Age: 10 years. Surgical resection status: Sub-total resection. Interval 1–2: ∼7 months. Interval 2–3: ∼12 months. Interval 3–4: ∼6 months*.*”*

### 2.4 Model Architectures

#### Time Encoding

For each patient trajectory, the inter-scan intervals are formatted into an ordered list [Δt_1_.….Δt_T_] where Δt_1_=0 and subsequent values represent the cumulative months between scans. The Δt list is then passed through a learnable linear projection to map into the 32-dimensional feature representation.

#### Causal Self-Attention

Causal self-attention is used across all three model architectures to preserve the temporal ordering of longitudinal MRI scans. Unlike standard self-attention, the causal self-attention restricts each timepoint t to attend only to itself and earlier scans in the sequence. This ensures chronological consistency in information progression,

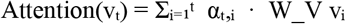

Where W… V… etc (define equation). This temporal masking enforces chronological correlation, allowing the transformer to encode how radiographic changes unfold over time while preventing information leakage from future observations.

#### Vision-Only Model

The vision-only architecture processes longitudinal MRI scans using a frozen BrainIAC [24] encoder, a foundation model pretrained on large-scale, heterogeneous brain MRI data to learn generalizable representations across sequences and institutions. BrainIAC was selected to provide a robust, task-agnostic visual representation to mitigate overfitting and improve generalization in limited-cohort prognostic settings. Each scan s_t is encoded into a visual token v_t = f_θ(s_t). The sequence{v_1_, v_2_, …, v_T} is then passed through a causal self-attention module to capture temporal dependencies. To incorporate timing, the learned 32-dimensional embedding u_i_ is concatenated with the visual token v_i_ to form ẑ_i_ = [v_i_ ∥u_i_], which is passed into the causal attention transformer. The final pooled representation z_vis is passed to a classifier to predict 1-year recurrence.

#### Clinical-Vision Model

The clinical-vision model extends the vision-only pipeline by incorporating structured clinical variables (sex, age at diagnosis, resection status) and the time interval list. Frozen BrainIAC encoder is followed by causal self-attention to generate a longitudinal imaging embedding z_vis similar to the vision-only model. In parallel, each clinical variable is encoded through a MLP. All clinical embeddings are concatenated to form a unified 128-dimensional clinical feature vector c. The final multimodal representation is obtained by concatenating imaging and clinical embeddings:

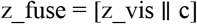

This fused representation is passed to a classifier, enabling the model to jointly learn over longitudinal imaging and clinical covariates.

#### Vision-Language Model (VLM)

The VLM introduces a text-based multimodal fusion for modeling longitudinal images and clinical context (Figure 1a). Structured clinical variables and time intervals are converted into a natural-language prompt describing the patient’s surveillance history. The frozen biomedical language encoder (Bioformer-8L) processes this prompt to produce contextualized text embedding (CLS token). Bioformer-8L was chosen for its lightweight design and efficiency, though the architecture is model-agnostic and can incorporate alternative biomedical SLMs or LLMs depending on the clinical content and complexity of the input text. In parallel, BrainIAC encodes each MRI scan, generating visual tokens v_1_,…,v__T_, which are then passed through causal self-attention to obtain the longitudinal imaging representation z_vis. Multimodal alignment is performed using cross-attention layers, which process imaging features as Key and Value and text representation as Query. By embedding Δt explicitly within the textual prompt, the model learns to represent temporal structure as part of the patient’s clinical narrative, enabling alignment between radiographic evolution and clinically meaningful temporal cues. The final fused representation is passed through a classifier to predict 1-year recurrence from the most recent scan.

### 2.5 Training objective, Evaluation and External Validation

All models were trained end-to-end using a binary cross-entropy objective. Subject-level stratification was enforced to prevent leakage of longitudinal information across training, validation, and testing sets. Training employed the AdamW optimizer with cosine learning-rate scheduling and early stopping based on validation area under the receiver operating characteristic curve (AUROC).

Models were trained using the DFCI/BCH cohort, which was partitioned at the subject level into training/validation and holdout test sets using an 80:20 split. The holdout DFCI/BCH set served as the internal test cohort.

The model performance was subsequently evaluated across four independent cohorts. The primary endpoint was prediction of 1-year recurrence using the most recent longitudinal scan prior to the outcome-defining event. Performance was assessed on the internal DFCI/BCH holdout test set as well as three external validation cohorts: RadART, CBTN, and PNOC008.

#### DeLong test methodology

The DeLong test is a nonparametric approach for comparing correlated ROC curves that provides an estimated variance-covariance matrix for AUCs and an asymptotic z-test of their difference[26-28]. In our study, predictions from two models were first aligned at the patient level using a shared identifier; for each pair of models, we constructed matched vectors of ground-truth labels *y*. We used the fast DeLong algorithm, which computes the AUC difference, its variance, the corresponded z-statistic, and a two-sided p-value using the normal approximation[26-28].

## 3 Results

### 3.1 Vision Language Modeling improves Recurrence prediction

Across the four independent external test cohorts (DFCI/BCH, CBTN, RadART, and PNOC008), the proposed VLM consistently outperformed both the clinical-vision and vision-only baselines. In the DFCI/BCH cohort, VLM achieved the highest discrimination (AUC = 0.94), exceeding the clinical-vision model (AUC = 0.91) and substantially outperforming the vision-only model (AUC = 0.69). Performance delta was most pronounced in the CBTN cohort, where VLM improved over clinical-vision (0.76 vs 0.58; p < 0.05) and vision-only (0.76 vs 0.53; p < 0.05). Similar improvements were observed in RadART (0.79 vs 0.72; p < 0.05). In the PNOC008 cohort, VLM again achieved the highest AUC (0.81), outperforming both baselines (clinical-vision: 0.58; vision-only: 0.56).

To further evaluate performance by tumor grade, we pooled cohorts into a low-grade glioma (LGG) group (DFCI/BCH + CBTN) and a high-grade glioma (HGG) group (RadART + PNOC008). In the LGG pooled set, VLM achieved an AUC of 0.75, significantly outperforming clinical-vision (0.70; p < 0.01) and vision-only (0.63; p < 0.01). In the HGG pooled set, VLM again showed the highest discrimination (0.79 vs 0.70 clinical-vision and 0.62 vision-only)..

### 3.2 Clinical Variables Masking Ablation

To quantify the influence of each clinical attribute, we performed structured masking of individual and pairwise variables for both the Clinical-Vision and VLM pipelines. Across all four cohorts, ablating surgical resection status, age, sex, or time-interval information resulted in reductions in performance. For the VLM, surgical resection status and inter-scan time intervals emerged as the most clinically informative features; masking either alone produced substantial declines, and masking them in combination (resection+time or age+time) led to the steeper decrements in predictive performance across datasets (CBTN: AUC 0.76 to 0.60; DFCI/BCH: 0.93 to 0.78; PNOC008: 0.81 to 0.65; RadART: 0.79 to 0.68). In contrast, masking sex yielded only modest reductions, indicating a smaller prognostic contribution.

The Clinical-Vision baseline demonstrated similar directional patterns, with resection status and time information driving the largest performance decrements. However, the magnitude of decline was consistently smaller and the overall performance lower than that of the VLM. Our ablation results demonstrate that temporal information and surgical resection status encode clinically meaningful information that allows better prognostication.

## 4 Conclusion

We introduced a vision–language framework for longitudinal MRI-based recurrence prediction in pediatric brain tumors. By expressing clinical variables and scan interval information as structured text and aligning them with imaging trajectories through cross-modal attention, our proposed model architecture provides a flexible way to incorporate heterogeneous clinical context without modifying the underlying architecture. Across four independent cohorts, the proposed VLM approach consistently improved predictive accuracy over vision-only and clinical-fusion architectures. The performance improvement was consistent in both low-grade and high-grade glioma subgroups. Our masking ablation experiments showed that time interval between scans and surgical resection status encode information that are the most informative of recurrence. Masking these variables led to systematic decreases in performance across all datasets, with the steepest drops observed when time intervals and resection status were removed in combination.

Although developed for pediatric neuro-oncology, the architecture is broadly applicable to other longitudinal prediction problems in oncology and neurology, where disease longitudinal imaging is acquired and along with the clinical assessments. Future work will evaluate the approach on additional clinical endpoints, assess performance in prospective surveillance validation, and explore how richer textual inputs, such as radiology reports or treatment summaries, can further strengthen temporal risk modeling.

## Data Availability

All data supporting the findings described in this manuscript are available in the article, in the Supplementary Information, and from the corresponding author upon request. The CBTN data is available upon request at https://cbtn.org/. The DFCI/BCH, PNOC, and RadART brain tumor dataset contains private hospital data that is controlled due to privacy concerns. Access to the derived dataset will be considered upon request to the corresponding author (Benjamin H. Kann, M.D., timeframe for response 2 weeks).

